# Exposure assessment for airborne transmission of SARS-CoV-2 via breathing, speaking, coughing and sneezing

**DOI:** 10.1101/2020.07.02.20144832

**Authors:** Jack Schijven, Lucie C. Vermeulen, Arno Swart, Adam Meijer, Erwin Duizer, Ana Maria de Roda Husman

## Abstract

**Background:** Evidence for indoor airborne transmission of SARS-CoV-2 is accumulating. If SARS-CoV-2 also spreads via aerosols, this has implications for measures taken to limit transmission.

**Objectives:** The aim of this study is to assess exposure to airborne SARS-CoV-2 particles from breathing, speaking, coughing and sneezing in an indoor environment.

**Methods:** An exposure assessment model was developed to estimate numbers of SARS-CoV-2 particles in aerosol droplets, expelled during breathing, speaking, coughing and sneezing by an infected person in an unventilated indoor environment, and subsequent inhalation by one or more persons. Scenarios encompass a range of virus concentrations, room sizes and exposure times.

**Results:** The calculated total volume of expelled aerosol droplets was highest for a sneeze, followed by a cough and speaking for 20 minutes, and lastly breathing for 20 minutes. A few to as much as tens of millions of virus particles were expelled. Exposure probability strongly depends on the viral concentration in mucus, as well as on the scenario. Exposure probabilities were generally below 1% at a virus concentration in mucus below 10^5^ per mL for all scenarios, increasing steeply at different higher concentrations. According to nose / throat swab data collected from patients, 75%, 50% and 5% of infected individuals carry an estimated number of SARS-CoV-2 per mL mucus of at least 10^5^, 10^6^ and 10^8^, respectively.

**Discussion:** Exposure to SARS-CoV-2 via aerosols generated during breathing, speaking, coughing and sneezing in an unventilated indoor environment is possible. This study forms a basis to estimate probabilities of exposure to SARS-Cov-2 by airborne transmission in indoor spaces. As long as it is uncertain what fraction of the airborne virus particles is infectious and as long as a dose response relation is lacking, it is recommended to be precautious.

## 1. Introduction

The fate of droplets in the air is mostly determined by their size; larger droplets deposit quickly while smaller droplets can stay airborne for longer periods, in so-called aerosols (a suspension of droplets in air). Aerosols can arise from ‘violent expiratory events’ such as coughing and sneezing (Bourouiba et al. 2014), but also from breathing and speech (Asadi et al. 2019; Leung et al. 2020). A dry cough is a predominant symptom of coronavirus (CoV) disease 2019 (COVID-19) (Wang et al. 2020). The WHO defines a cut-off of 5 µm to distinguish airborne (<5 µm) from droplet (> 5 µm) transmission (World Health Organization 2014, 2020). The existence of a cut-off is contested, other organizations use different terminology and cut-off values (Kohanski et al. 2020; Tellier et al. 2019). Infectious particles <5 µm can penetrate more deeply into the lungs, while larger particles most likely impact the upper airways (Gralton et al. 2011; Tellier et al. 2019). While it is true that the large majority of the volume of fluids that is expelled during, for example, coughing and sneezing, is in droplets that deposit quickly, this does not imply that airborne transmission is highly unlikely (Nicas et al. 2005). Furthermore, research suggests that the cut-off size of droplets (aerodynamic diameter) which deposit quickly is higher than 5 µm, and not static but dependent on a number of factors, such as relative humidity (Liu et al. 2017). In the absence of turbulence, droplets with an initial diameter larger than 80 µm will be deposited on the floor from an initial height of 2 m at a distance away from the mouth of around 1 m (Liu et al. 2017). The droplet with an initial diameter of 60 µm can reach about 4 m, with a size of 0.32 times its initial diameter at a relative humidity (RH) of 0%, whereas it can travel a distance of 1.85 m at a RH of 90% due to its larger droplet size of 0.43 its initial diameter (Liu et al. 2017). In the case of turbulence, even initially larger particles could likely travel even further. Therefore, airborne and droplet transmission occur on a continuum, and airborne transmission can potentially occur in the size fraction of all particles less than about 60 µm (Gralton et al. 2011; Kohanski et al. 2020; Tellier et al. 2019).

Droplets and aerosols can harbour pathogens such as bacteria e.g. *Coxiella burnettii*, and viruses such as influenza viruses (Milton et al. 2013; Stein et al. 2005). Evidence exists for airborne (bioaerosol) transmission of multiple viral respiratory diseases, including SARS, MERS and influenza (Adhikari et al. 2019; Kulkarni et al. 2016; Weber and Stilianakis 2008; Yu et al. 2004; Zhang et al. 2013). Airborne transmission has also been suggested as probable for SARS-CoV-2 (Anderson et al. 2020; Asadi et al. 2020; Chia et al. 2020; Correia et al. 2020; Li et al. 2020; Morawska and Cao 2020; Richard et al. 2020; Setti et al. 2020; Shen et al. 2020; Stadnytskyi et al. 2020; Wang and Du 2020; Yao et al. 2020), although other studies contest this and suggest airborne transmission does not take place (Xu et al. 2020).

SARS-CoV-2 has been observed to be remarkably stable in aerosols generated under laboratory conditions (Fears et al. 2020), with little decline in infectivity after 16 hours of aerosol suspension. Similarly, van Doremalen et al. (2020) also found that SARS-CoV-2 remained viable for hours in experimentally generated aerosols (reduction in infectious virus particles from 3100 to 500 per litre air in 3 hours).

There is much discussion about the potential for airborne transmission of SARS-CoV-2. The World Health Organization so far maintains that COVID-19 is not airborne (https://who.africa-newsroom.com/press/coronavirus-fact-check-covid19-is-not-airborne). Eissenberg et al. (2020) discuss the evidence cited by WHO and conclude that it remains prudent to consider airborne transmission of COVID-19 as an explanation for the rapid spread of the virus. If SARS-CoV-2 also spreads via airborne transmission, this has implications for the measures that are being taken to limit transmission, such as advice to keep a certain distance from other people. If SARS-CoV-2 is also airborne, transmission would be plausible beyond the often advised 1.5 meters.

The aim of this study is to assess exposure to airborne SARS-CoV-2 particles from breathing, speaking, coughing and sneezing in an indoor environment. Figure 1 shows a schematic overview of the processes modelled in this study. The exposure assessment entailed estimating the numbers of SARS-CoV-2 particles in aerosol droplets, expelled during breathing, speaking, coughing and sneezing by an infected person in an unventilated indoor environment, and subsequent inhalation by one or more persons in that environment. Exposure assessment is part of a Quantitative Microbial Risk Assessment (QMRA), in which exposure assessment is followed by a risk characterisation. In risk characterisation, estimates of risk of infection and/or illness are made using dose response relations (Haas et al. 1999). In this study, risks of infection/illness were not estimated because dose response relation data are lacking for SARS-CoV-2. Literature and laboratory data on SARS-CoV-2 virus concentrations from nasopharyngeal and oropharyngeal swab samples (hereafter: nasal and throat), numbers of expelled aerosol droplets and their size distributions were used as input for the model calculations. This study focused on modelling airborne transmission and thus did not model larger droplet transmission, e.g. the probability of coming into contact with large droplets that fall directly on mucosa or surfaces.

**Figure 1.**
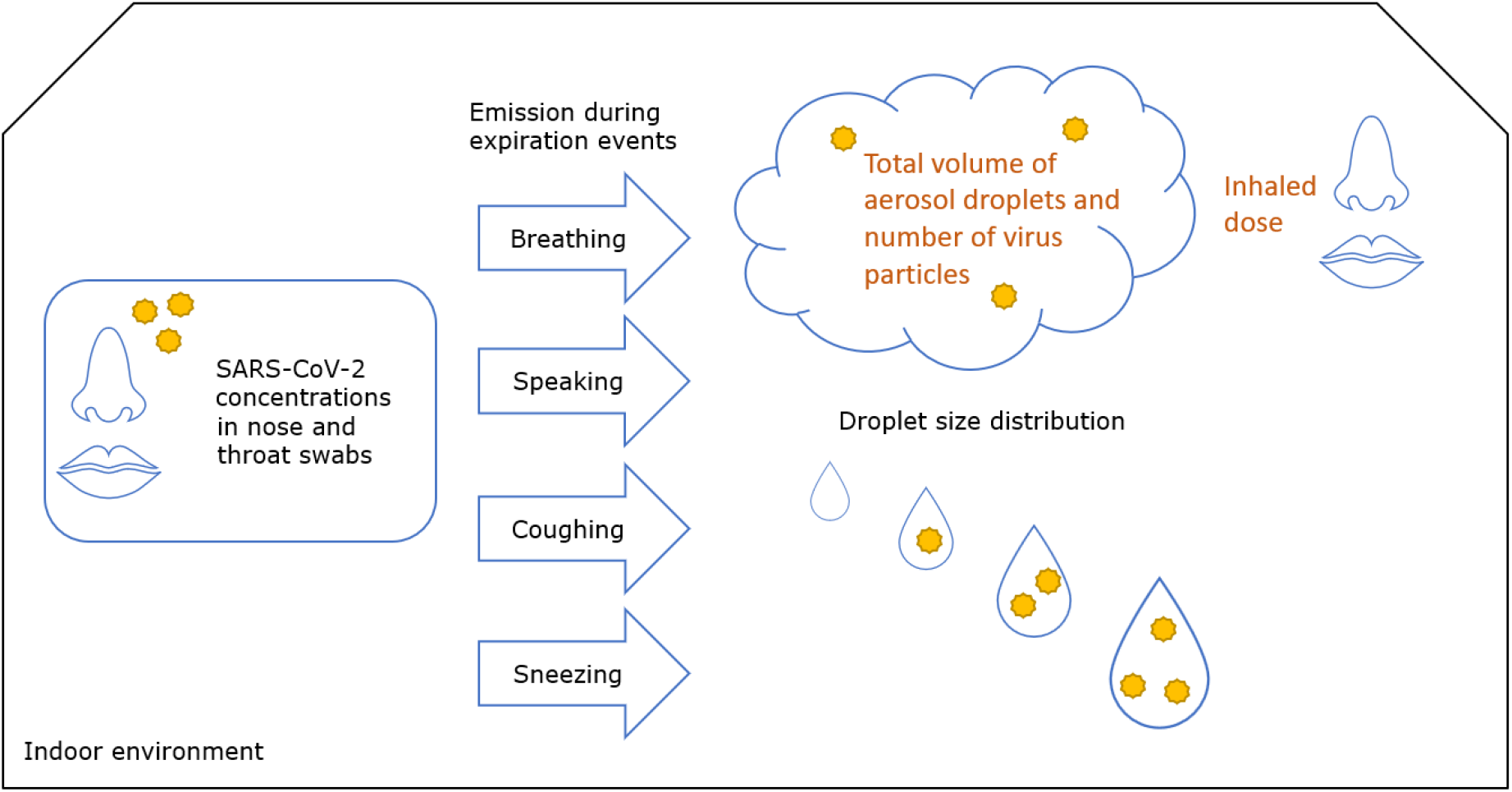
Overview of the processes modelled in this study.

## 2. Methods

### 2.1 Scenarios

Table 1 shows the scenarios that were applied. All scenarios consist of two parts. The first part is the expelling part, where an infected, (a)symptomatic person enters a room and stays there for 20 minutes. Two differently sized rooms were considered, one room with the dimensions of 12 × 2.55 × 3.05 m^3^ = 93 330 litres, which corresponds with the dimensions of a Dutch public transport bus with two axles (GVB https://over.gvb.nl/ov-in-amsterdam/feiten-en-cijfers/bus-in-cijfers/), and the other room with dimensions of 15 × 15 × 3 m^3^ = 675 000 litres. Such a room may be a small restaurant, fitness room, or meeting room, etcetera. For convenience the two rooms are denoted as bus and room. The infected person expels virus particles through 20 minutes of breathing, 20 minutes of speaking, one cough, or one sneeze. The bus and room are not ventilated, and it is assumed that the virus particles that are contained in aerosol droplets are evenly distributed in the rooms. In the second part of the scenarios, the exposure part, one or more persons are exposed to the virus contained in the aerosol droplets. In the case of the bus, one or thirty other persons enter for 20 minutes and in the case of the room, ten persons enter for 1 hour or 4 hours.

**Table 1.** Selected scenarios for which aerosol exposure was quantified.

| *Expelling of virus by one infected person* |  |  |
| --- | --- | --- |
| Abbreviation | Source | Reference of size distribution data |
| Breathe | 20 minutes breathing | Fabian et al. (2011) |
| Speak-Lo | 20 minutes speaking | Asadi (2019) |
| Speak-Hi | 20 minutes speaking | Duguid (1946) |
| Cough-Lo | One cough | Lindsley et al. (2012) |
| Cough-Hi | One cough | Duguid (1946) |
| Sneeze-Lo | One sneeze | Gerone et al. (1966) |
| Sneeze-Hi | One sneeze | Duguid (1946) |
| *Exposure of one or more uninfected persons* |  |  |
| One person in a bus for 20 minutes |  |  |
| Thirty persons in a bus for 20 minutes |  |  |
| Ten persons in a room for one hour |  |  |
| Ten persons in a room for four hours |  |  |

One reason for using different scenarios for expelling virus was because of inconsistencies in the literature regarding the number and size distribution of the aerosol droplets from speaking, coughing and sneezing. For speaking, two scenarios, low and high, include the data from size distributions from Asadi (2019), reading an English passage aloud at intermediate loudness (85 dB), and Duguid (1946), speaking loudly. Similarly, two scenarios encompass the data from size distributions for coughing, denoted as low and high from Lindsley et al. (2012) and Duguid (1946), respectively. Likewise, two scenarios, low and high, encompass the data from the size distributions in a sneeze as reported by Gerone at al. (1966) and Duguid (1946), respectively. In the selection of data from literature, studies concerning bacterial infections were not included. Size distribution data for breathing were based on the study by Fabian et al. (2011). Fabian et al. (2011) concerned human rhinovirus-infected subjects, Gerone (1966) concerned coxsackie virus-infected subjects, Lindsley et al. (2012) concerned influenza virus-infected subjects, and all other studies listed in Table 1 concerned studies on healthy individuals. The Supplementary material contains more discussion on the choice of the size distribution data.

Exposure assessment was conducted with each value of the following decimal range of the virus concentration in mucus: 10^3^, 10^4^, 10^5^, 10^6^, 10^7^, 10^8^, 10^9^, 10^11^ virus particles per mL. This range in virus concentrations as estimated from viral RNA detection reflects those observed in nasal and throat swabs (own data, 2020) (Zou et al. 2020).

### 2.2 Exposure assessment model

#### Virus concentration

Viral concentration from throat and nasal swabs were determined for the first 729 SARS-CoV-2 E-gen positive diagnostic samples sent in from municipal health services and hospitals to the National Institute for Public Health and the Environment of the Netherlands (RIVM) for COVID-19 diagnosis, analysed as decribed (Corman et al. 2020). For quantification, the RdRp gene fragment was used. These data and data from Zou and co-workers (Zou et al. 2020) were fitted to a left-censored lognormal distribution using a Bayesian fit in RStan (R Core Team 2019; Stan Development Team 2020), in which the logarithm of the virus concentration was assumed to be normally distributed and from the onset of symptoms to decrease in time:

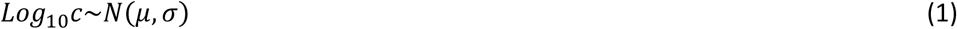

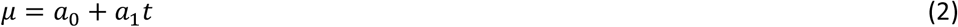

where *c* is the virus concentration [numbers per mL mucus] with mean *µ*, standard deviation *σ*, coefficients *a*_*0*_ and *a*_*1*_ and time *t* [days] from onset symptoms. Note that, in the Zou-data, a distinction between the data from nasal and throat swabs was made, but this was not possible in the RIVM-data where most samples were combined nose-throat swabs. SARS-CoV-2 concentrations are determined as Ct values, which are inversely related to viral RNA copy number. Viral RNA copy number in mucus was estimated from Ct values, as detailed in Supplementary material S1.

#### Total volume of aerosol droplets

The total initial volume of aerosol droplets per cough, sneeze, 20 minutes speaking and 20 minutes breathing was calculated from the number of aerosol droplets and their size distribution. Following Liu et al. (2017), in collecting literature data of the size distribution of expelled droplets by breathing, speaking, coughing and sneezing, expelled droplets smaller than 60 µm were considered, when measured directly in front of the mouth, assuming little evaporation had happened. Liu et al. (2017) reported that the droplet nuclei size at a relative humidity of 90% (25°C) could be 30% larger than the size of the same droplet at a relative humidity of less than 67.3% (25°C). In the case of a distance of about 0.5 m or more, the size distribution of droplets of 20 µm or less was considered, and for these droplets, their initial size distribution (at the point of leaving the mouth) was estimated by multiplying their diameter by a factor of three, to correct for evaporation (Liu et al. 2017).

For the total aerosol droplet volume by breathing, the data reported by Fabian et al. (2011) were used. Fabian et al. (2011) reported a box-whisker chart of the logarithm of the number of aerosol droplets per litre of exhaled air for six aerosol droplet size classes from 0.3 µm to >10 µm. For each size class the average diameter was used, for >10 µm the value of 15 µm was chosen. From box-whisker chart mean and quantile values were extracted to estimate mean and standard deviation of the logarithm of the number of aerosol droplets per litre of exhaled air for each of the six aerosol droplet size classes. It was realised that a correlation between the numbers of droplets expelled in the different size classes may exist that was not apparent from this chart (e.g. a subject that expels above average droplets in one size class is perhaps likely to also expel above average in other size classes). The total volume of aerosol droplets per minute of exhaled air, *v*_*br*_ was calculated as follows:

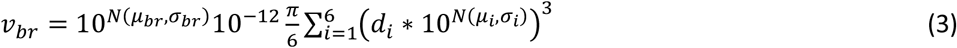

where *d*_*i*_ is the diameter [µm] of aerosol droplets in the *i*-th class of six aerosol droplet diameters, of which the logarithm of their concentration in air is normally distributed with mean *µ*_*i*_ and standard deviation *σ*_*i*_. The volumes of each aerosol droplet size were summed, then converted from µm^3^ to millilitres by a scaling factor of 10^−12^ and multiplied by the tidal breathing rate that is normally distributed on log-scale with mean *µ*_*br*_ and standard deviation *σ*_*br*_ (Table 2).

**Table 2.**
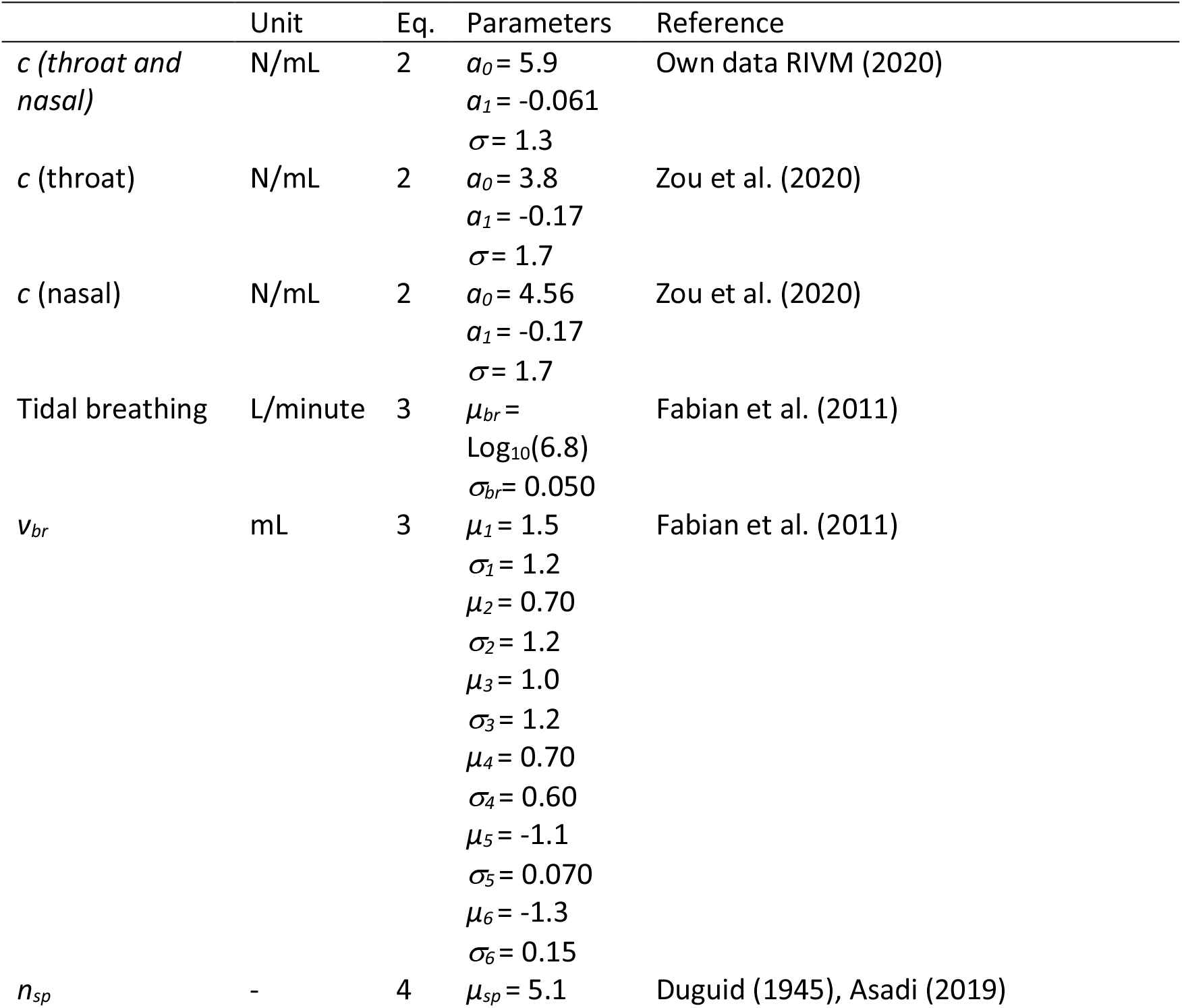
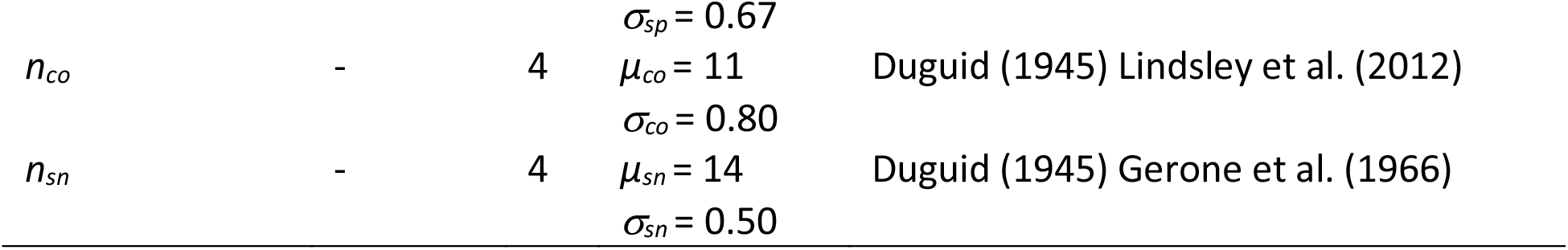
Parameter values

The numbers of expelled aerosol droplets *n*_*sp*,*co*,*sn*_ during speaking, coughing or sneezing, respectively, were assumed to be lognormally distributed:

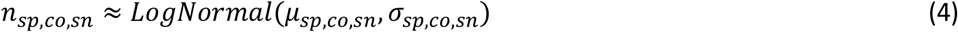

The ≈ sign denotes rounding to integer values. Values of parameters *µ*_*sp*_ and *σ*_*sp*_ are given in Table 2 and represent the range of numbers of aerosol droplets as reported by Duguid (1945) and Asadi et al. (2019) per litre and per minute of speaking loudly. In the case of Asadi et al. (2019), these numbers needed to be scaled to the volume of air a person exhales during speaking a minute using the values for tidal breathing increased by 13.5% (Bunn and Mead 1971). Duguid reported data for subjects counting from 1 to 100, it was assumed this represented an observed speaking time of 1.5 minutes. Values of parameters *µ*_*co*_ and *σ*_*co*_ represent the range of numbers of coughed aerosol droplets as reported by Duguid (1945) and Lindsley et al. (2012). Values of parameters *µ*_*sn*_ and *σ*_*sn*_ represent the range of numbers of sneezed aerosol droplets as reported by Duguid (1945) and Gerone et al. (1966).

For each study, the total volume of aerosol droplets per 20 minutes of speaking, per cough and per sneeze, *v*_*sp*,*co*,*sn*_ [milliltres] was calculated by summing *n*_*sp*,*co*,*sn*_ samples of each diameter data set as follows:

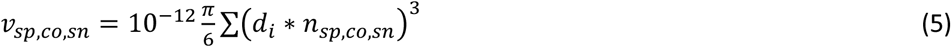

The size distributions or aerosol droplets expelled by speaking were from Duguid (1946) and Asadi et al. (2019). Asadi et al. counted particles between 0.5 – 20 µm with highest counts around 1 µm, while Duguid reported underestimation of particles less than about 1 µm with highest counts around 2-4 µm. The size distributions or aerosol droplets expelled by coughing where taken from Lindsley et al. (2012) and Duguid (1946). Both reported counts of particles for the various aerosol droplet diameters. The size distributions or aerosol droplets expelled by sneezing where taken from Gerone et al. (1966) and Duguid (1946) Gerone et al. (1966) did not count particles larger than 15 µm, and Duguid reported underestimation of particles less than about 1 µm.

See Supplementary material S2 for more detailed discussion on data selection, and the counted aerosol droplet diameter ranges, method of counting and subjects investigated in the selected studies.

#### Dose and exposure probability

It was assumed that the expelled aerosol droplets were instantaneously evenly dispersed in the room. The dose *D* was computed as follows:

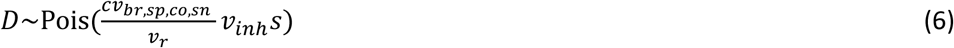

where *v*_*r*_ is the volume of the room [litres], and *v*_*inh*_ is the inhaled volume of an exposed person during 20 minutes using the same tidal breath rate as in equation (3), *s* is a sensitivity factor.

In equation (6), *s*c* can be interpreted as a change in *c* by factor *s*. Similarly, *s*v*_*br*,*sp*,*co*,*sn*_, *s/v*_*r*_ and *s*v*_*inh*_ can be interpreted as a change in *v*_*br*,*sp*,*co*,*sn*_, 1/*v*_*r*_ and *v*_*inh*_. In other words, any change in these variable or combination of these variables by factor *s* has the same effect on the dose and probability of exposure. Factor *s* can also be interpreted as the number of exposed persons, or the number of persons expelling virus. Also note that a factor *s* change in *v*_*inh*_ can be due to a change in exposure time.

In the scenario with one exposed person in a bus for 20 minutes, *s*= 1. This is the reference scenario. In the scenario with 30 exposed person in a bus for 20 minutes, *s*=30. In the larger room with 10 persons for 1 hour, *s*=1/7*10*3=4, and in that room for 4 hours, *s*=16. For the sensitivity analyses two extra scenarios were added with factor *s*=0.1 and *s*=100.

Finally, the probability of exposure *P*_*exp*_ was computed as one minus the Poisson probability of exposure to zero particles:

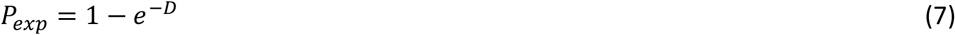

All volumes of aerosol droplets, the dose and probability of exposure were computed by drawing 10 000 Monte Carlo samples. All computations were conducted using Mathematica (Wolfram Research Inc. 2019).

Because short time frames are considered, and SARS-CoV-2 has been observed to remain infectious in aerosols for hours (Fears et al. 2020; van Doremalen et al. 2020), decay over time is not modelled in this study. The exposure assessment will, in that regard, be conservative.

## 3. Results

### 3.1 Parameter values

Table 2 shows the parameter values as obtained for the different equations. For speaking, coughing and sneezing, the particle size distribution could not be described properly by a lognormal, gamma, or normal distribution as indicated by an Anderson-Darling test. Therefore, Monte Carlo simulation entailed direct random sampling from the size distribution data.

### 3.2 Exhaled volumes of aerosolized droplets

The total volume of expelled aerosolized droplets as calculated for the different scenarios for breathing, speaking, coughing and sneezing (eqs. 3 and 5) is shown in Figure 2. Variation is observed both within and between scenarios. For sneezing there is about one order of magnitude difference between the low and high scenarios (mean from 5 000 to 30 000 picolitres per sneeze). For coughing there is about two orders of magnitude difference between the low and high scenario (mean from 30 to 4 000 picolitres per cough). For speaking, the two scenarios are similar (mean from 200 to 300 picolitre per 20 minutes). The breathing scenario varies about seven orders of magnitude (mean 6, range from 0.004 to 30 000 picolitres per 20 minutes), the speaking and coughing scenarios vary about three orders of magnitude and the sneezing scenarios vary about two orders of magnitude. The total aerosol volume from speaking loudly for 20 minutes is in between that of the low and high scenarios from one cough.

**Figure 2.**
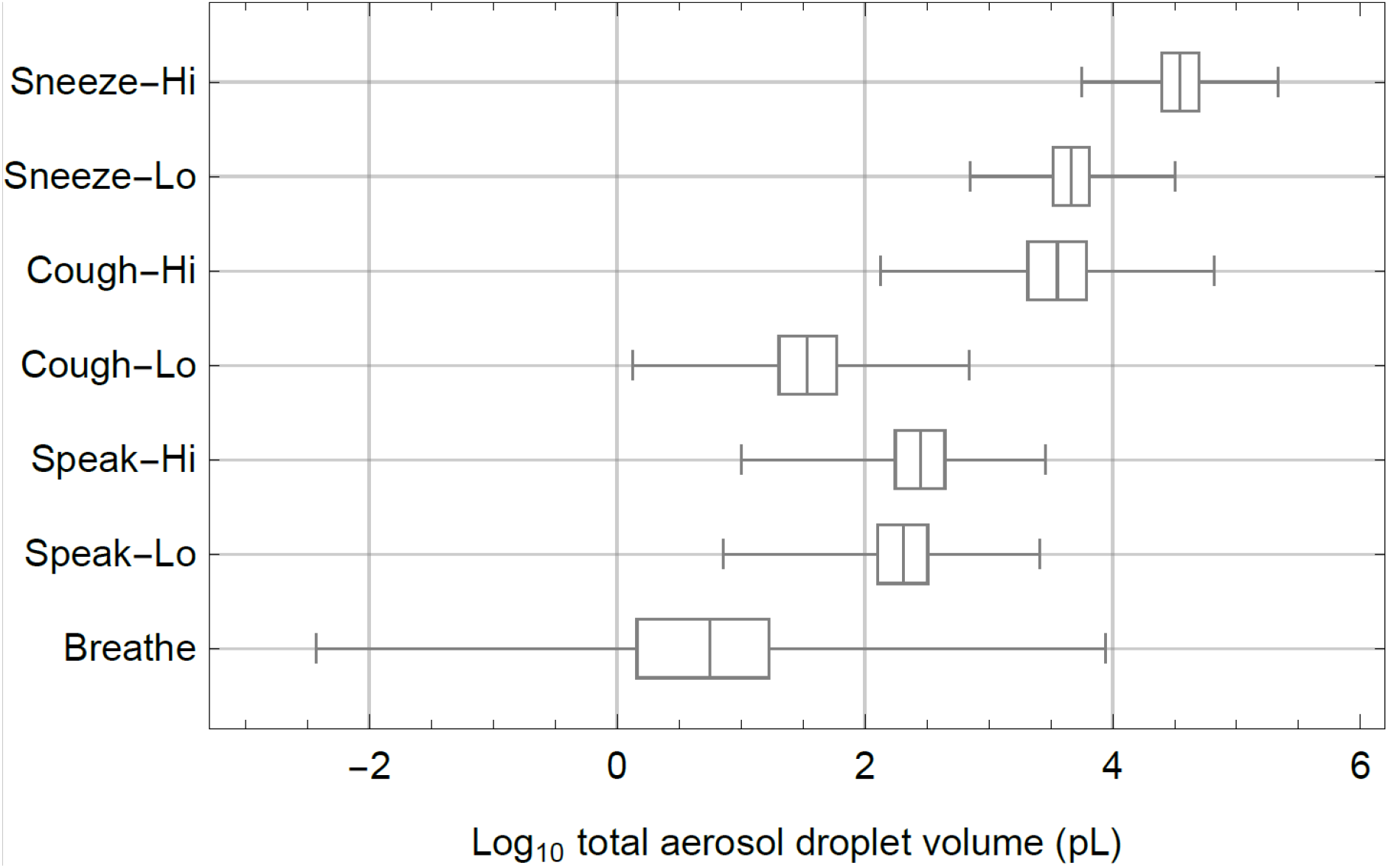
Box-whisker chart of the log_10_ of total aerosol droplet volumes (pL=picolitres) that are expelled in each scenario (Table 1), showing median values, quartiles (boxes) en minimum and maximum values (whiskers). Volumes in pL/20 minutes for breathing and speaking and in pL per cough and per sneeze.

### 3.3 Viral concentration data from swabs

Observed SARS-CoV-2 concentrations in mucus spanned a wide range, from 10^2^ to 10^11^ copies / mL (corresponding to a range of Ct values from 40 to 10.5). Observed concentrations in mucus were on average 2 orders of magnitude higher for the RIVM data (∼10^6^ copies / mL) than in the Zou data (∼10^4^ copies / mL, Figure 3 bottom panels). SARS-CoV-2 virus concentrations from the nasal swabs of Zou et al. (2020) were, on average, higher than those from throat wabs, and both decreased over time (days since symptom onset) (Supplementary material S1). This decrease in concentration from the onset of symptoms was also observed in the own data. The bottom panels of Figure 3 show the probability densities of the virus concentrations in swabs from infected persons in parallel with the range of virus concentrations applied in the scenarios. This way it is possible to read the probability that a person expels virus at a certain concentration. It was chosen to show the probability densities instead of the raw data, as this allowed for including non-detects in the fits and gives a better view of the mean and 95% of the data.

**Figure 3.**
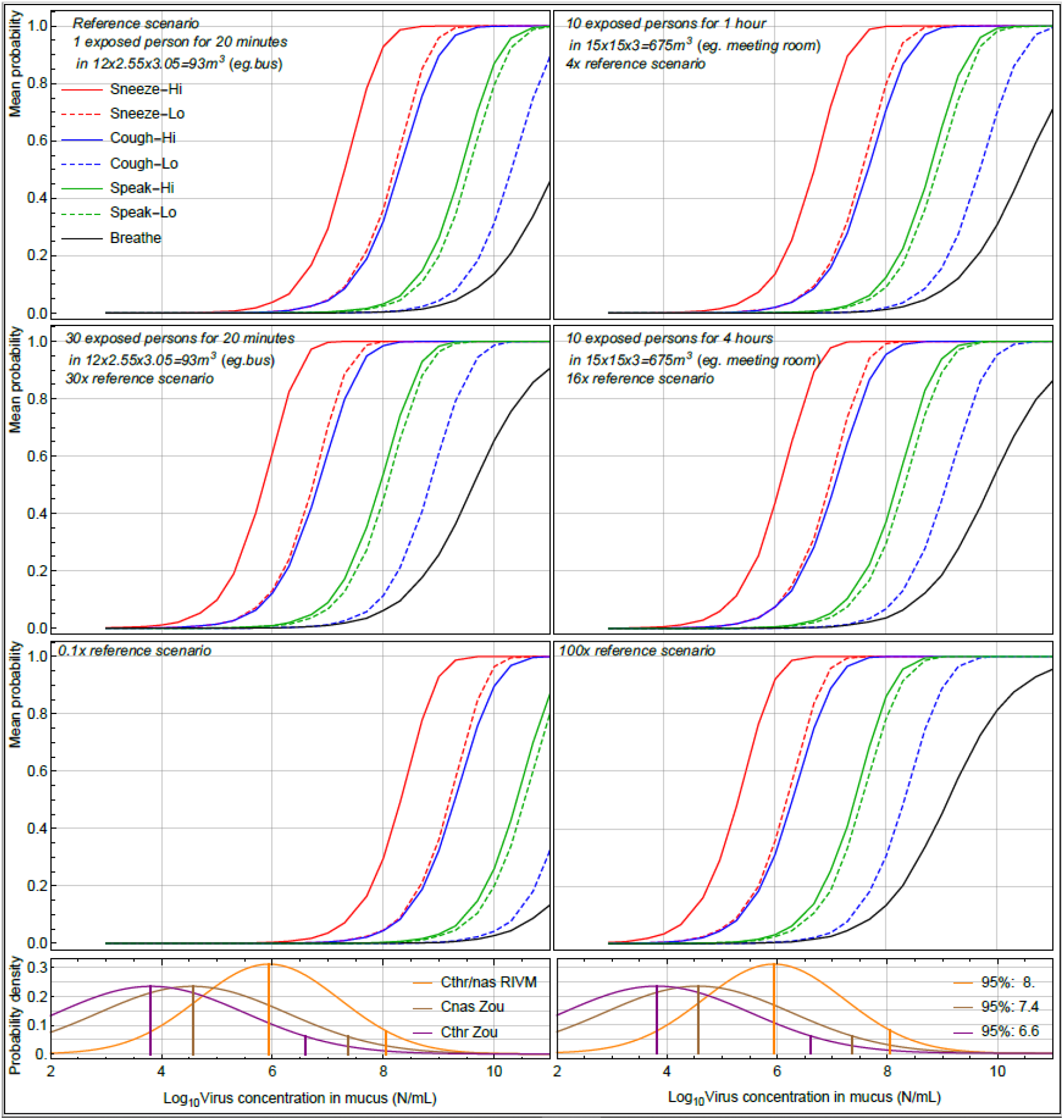
Mean probability of exposure to SARS-CoV-2 for different scenarios, in which one infected person was expelling virus by 20 minutes of breathing, 20 minutes of speaking, one cough or one sneeze. This happened in a room of 93 m^3^ or 675 m^3^ with exposed persons (1, 10 or 30 persons) for a certain amount of time (20 minutes, 1 hour, 4 hours) as indicated. In the scenarios with multiple exposed persons, the shown probability is for at least one person of the group being exposed. Each mean exposure probability curve is for a given virus concentration in mucus. Below the exposure probability curves, probability densities of the virus concentration in nasal and throat swabs (Zou et al. 2020) and of nasal or throat swabs (own data, 2020) are given with mean and 95-percentiles indicated (vertical lines) to reflect the probability of a given virus concentration in mucus.

### 3.4 Numbers of expelled virus particles and dose

The number of viral particles that is expelled by an infected individual is dependent on the virus concentration and the volume expelled. At lower virus concentrations (e.g. <10^5^ copies / mL in the mucus), there is a large probability that no viruses are expelled, especially during breathing and speaking. At higher virus concentrations, the range of virus particles that are expelled varies hugely from a few to more than ten million virus particles. Corresponding concentrations of virus in the air of the room ranged from 10^−4^ to 10^2^ per litre of air. Due to the variability in expelled volume, the expelled numbers of virus particles vary about a factor of one hundred for a given virus concentration for breathing, and about a factor of ten for the other scenarios. The numbers of expelled virus particles correspond to the expelled droplet volumes and vary about a factor of ten between the low and high scenarios for speaking and sneezing, and about a factor of one hundred between the low and high coughing scenarios. The dose follows a similar pattern, but numbers of virus particles are about a factor of one thousand lower because the inhalation volume is about one thousandth of the volume of air in the bus. Graphical details on the numbers of expelled virus particles and the associated dose are given in the Supplementary material S3 (Figure S1).

### 3.5 Probability of exposure

Figure 3 summarizes the probability of exposure for all scenarios. The exposure probability can be read from the curves, for a certain virus concentration. Note that the figure shows the mean probabilities, and these probabilities vary one to two orders of magnitude, like the number of expelled virus particles. Generally, low probability of exposure is observed at a virus concentration below 10^5^ per mL for all scenarios, except the 100x reference scenario. Furthermore, of course directly related to the volume of expelled aerosol droplets (Figure 2), it is observed that the probability starts to increase steeply at different concentrations for each of the scenarios. Going from the high sneezing scenario to the low sneezing scenario and the high coughing scenario, then to both speaking scenarios, followed by the low coughing scenario, and finally, the breathing scenario, for each these steps, about a ten times higher virus concentration is required for the same exposure probability. Going to a scenario with more exposed persons, longer time for exposure, or smaller room, the probability of exposure curves all shift to the left.

For example, at a virus concentration of 10^6^ per mL mucus and in the high sneeze scenario, in the bus with one exposed person, figure 3 indicates a probability of exposure of about 4%. In the same scenario, but with 30 exposed persons, the probability of exposing at least one person amounts to 60%. In the larger room, with 10 exposed persons, the exposure probability is 14% and 41% for the 1-hour and 4-hours exposure times, respectively. In a full bus and nobody speaking, coughing or sneezing, the probability of exposure is about 6% when an infected person expelled virus at a concentration of 10^8^ per mL. And if in that bus an infected person would sneeze heavily (the high-sneeze scenario), at a virus concentration of 10^8^ per mL, probability of exposure of at least one persons would be equal to one. At a concentration of 10^6^, this probability is 60%.

Visually comparing with the probability density distributions of the measured virus concentrations in the bottom panels, (own data, 2020) (Zou et al. 2020) one can get an impression of the probability of the corresponding virus concentration occurring. For example, following the RIVM data, a virus concentration of 10^8^ per mL or more in the mucus occurs in about 5% of infected individuals.

## 4 Discussion

The probability of exposure of at least one person to SARS-CoV-2 particles contained in aerosol droplets that were expelled by an infected person was estimated in various scenarios wherein the infected person was expelling virus by breathing, speaking, coughing or sneezing. An important assumption was that the initial aerosol droplets have the same virus concentration as measured in mucus from nasal and throat swab samples, and that the numbers of virus particles in mucus were evenly distributed. Moreover, it was assumed that the expelled aerosol droplets were instantaneously and evenly distributed in the air of the room. For the sake of simplicity for this first study, it was assumed that an infected person leaves the room before one or more susceptible persons enter. In further work, this could evolve to a dynamic model with rates at which virus is expelled and inhaled, as was done by Buonanno et al. (2020). Although the approach is similar, our results cannot directly be compared to the work by Buonanno et al., as they presented calculated risks for very specific scenarios.

The range of observed SARS-CoV-2 concentration in swab samples of 10^2^ – 10^11^ RNA copies / mL is similar to the range found in another Dutch study by van Kampen et al. (2020), who observed a range of 10^3^ – 10^10^ RNA copies / mL. The calculated range of viral concentrations in the air (from 10^−4^ to 10^2^ per litre of air, section 3.4) does encompass the values of observed airborne SARS-CoV-2 concentrations in hospital rooms with SARS-CoV-2 patients (Chia et al. 2020; Guo et al. 2020; Liu et al. 2020; Ong et al. 2020; Santarpia et al. 2020). Chia et al. (2020) observed total airborne SARS-CoV-2 concentrations ranging from 1.8 to 3.4 RNA copies/ L air. About half of the RNA was found in droplets sized 1 – 4 µm, and the other in half particles sized > 4 µm. Santarpia et al. (2020) similarly observed 0.98 to 8.7 copies / L air in hallway air samplers, and higher concentrations of 5.4 to 67 copies / L air with personal air samplers. From one of the samples, it was indicated that it contained culturable SARS-CoV-2, suggesting that not only RNA in inactivated virus was emitted to the air, but also infectious virus. Liu et al. (2020) measured air in COVID-19 patient areas, staff areas and public areas in two Wuhan hospitals. In all three areas positive samples were found with low concentrations, ranging from 10^−3^ to 10^−1^ RNA copies / L for patient areas, 10^−3^ to 4 × 10^−2^ for staff areas, and 10^−3^ to 10^−2^ for public areas. Highest SARS-CoV-2 concentrations were detected in droplet size ranges of 0.25 – 1 μm and >2.5 μm. Guo et al. (2020) similarly found air samples positive for SARS-CoV-2 RNA both in patient isolation wards but also the doctor’s office in a hospital. In contrast, Faridi et al. (2020) and Ong et al. (2020) were unable to detect SARS-CoV-2 RNA in the air in hospital wards. Ong et al. measured in well-ventilated airborne infection isolation rooms and their anterooms (12 air changes per hour). Although air samples were negative, the air outlet fan was found positive for SARS-CoV-2 RNA. This shows the importance of ventilation.

The assumption that expelled aerosol droplets are instantaneously and evenly distributed in the air of the room implies that there is an immediate dilution of the expelled virus concentration, which lowers its concentration in the air, but also spreads the virus. Obviously, dilution will not really occur instantaneously, it highly depends on the aerodynamics in the room. An exposed person directly in front of the infected person, or in a flow path of the contaminated air, may inhale a much larger dose than average. Clearly, air ventilation is very important. It may be surmised that in outdoor spaces exposure probability will be much less, due to much more dispersion and dilution.

In the exposure assessment, virus inactivation was not included. According to van Doremalen et al. (2020) the infectious virus concentration should have decreased 20 (8-30)% after 20 minutes, 47 (23-66)% after one hour and 92 (65-99)% after 4 hours. Including virus inactivation should require accounting for the virus decline time during exposure, on the other hand, in several situations, viruses are continuously expelled. Furthermore, it should be noted that it is unknown what fraction of airborne RNA-copies is infectious virus. Observational data on infectious virus in aerosols in various settings are needed to validate modelling efforts. No dose-response model is, as of yet, available for SARS-CoV-2. In future work, an existing dose-response model for a highly infectious agent analogous to the approach by Schijven et al. (2016) could be applied, to make a first estimate of risk to be infected, or the dose-response model for SARS 1 (Watanabe et al. 2010) could be taken as a starting point. Additionally, the probabililty of illness when infected is also unknown. For other respiratory viruses, e.g. adenovirus, the dose-response model quickly approaches a 100% infection rate at a dose of ∼10 viral particles, but 100% illness is not attained (Teunis et al. 2016). This should be further studied for SARS-CoV-2. The location of infection, in relation to the size of the droplets might also be of importance with respect to illness outcomes. Droplets <5 µm can penetrate more deeply into the lungs where SARS-CoV-2 can infect airway epithelium. SARS-CoV-2 entry factors were also found to be highly expressed in nasal epithelium meaning that infection can likely establish there as well (Sungnak et al. 2020). SARS-CoV-2 prevalence in the population of course influences the probability that an infected person is present in a bus or room, this was not accounted for in the model. Similarly, immunity in the population influences the probability that susceptible persons are present.

The current study was entirely focused on estimating probabilities of exposure to SARS-CoV-2 in aerosolized droplets that are small enough to be distributed in the air farther than 1.5 meter. From this study, the relative importance of aerosol and droplet transmission cannot be determined. To put this in perspective, Duguid et al (1946), also captured droplets with an initial diameter of more than 60 μm when speaking, coughing or sneezing, of which the total volume can be 3-5 orders of magnitude higher than the total volume of the smaller droplets <60 μm. However, for droplet transmission, different factors govern the probability of exposure, such as the probability of expelling droplets directly onto the mucosa of another person, hand hygiene, hand-mouth contact virus transfer rates, etcetera. It should be noted that, although transmission at short distances is commonly thought to be droplet transmission, Chen et al. (2020) found that the short-range airborne route actually dominates at most distances during both talking and coughing. Chen et al. studied airborne transmission in general, not specifically SARS-CoV-2 or another virus.

The highest numbers of viruses were expelled in aerosol droplets from a sneeze, followed by the high coughing scenario. Persons that are sneezing and/or coughing are advised by governments to stay at home. However, it is also reported that symptoms can start acutely and that the day before, and first days with, symptoms, the highest virus loads are found in naso-pharyngeal swabs (Kimball et al. 2020; Pan et al. 2020; Zou et al. 2020). Therefore, an occasional sneeze or cough could easily occur before one realizes one should have stayed at home. Somsen et al. (2020) found that sneezing only produces large drops and not so much aerosols, which is in contrast with our findings of the potential importance of sneezing. As coughing is a predominant symptom of COVID-19 (Wang et al. 2020), it can be envisioned that the scenario of a single cough is not realistic, and persons might cough many times in a row. This would render coughing perhaps more important than sneezing. Furthermore, singing, speaking loudly (speaking is commonly directed to other persons), breathing heavily because of intense activity, all may expel as many virus particles as coughing. Reports on high SARS-CoV-2 attack rates during choir practices exist, e.g. (Hamner et al. 2020). Singing is reportedly associated with tuberculosis outbreaks (Mangura et al. 1998), and Loudon and Roberts (1968) found the number of airborne droplet nuclei after singing to be higher than after talking, nearly as high as after coughing. Other studies also suggest that airborne transmission of infectious diseases is possible without coughing or sneezing, but simply from exhaled breath from individuals who barely show any symptoms (Asadi et al. 2019; Yan et al. 2018).

The observed droplet size distribution peak is different for studies measuring specifically in smaller size ranges (0.5 – 20 μm) (Asadi et al. 2019; Fabian et al. 2011), than in studies measuring in larger size ranges for both airborne and larger droplets (2-1000 μm). Studies measuring in the small range generally report the peak (highest numbers of particles) around 1 μm, while studies measuring in the larger range report the peak ∼15 μm (Chao et al. 2009; Duguid 1946). Difference in the sensitivity of the equipment for certain size classes might account for this discrepancy.

It is thought that for many infectious diseases that superspreaders drive a large part of the transmission in the population; for SARS 1 the 20% most infectious individuals were estimated to cause 80-90% of the transmission (Galvani and May 2005; Lloyd-Smith et al. 2005), and also for SARS-CoV-2 this has been suggested (Endo et al. 2020). This study underpins the idea of superspreading events via aerosols. High virus concentrations in mucus of infected individuals appear to be necessary for high probabilities of exposure for persons in the same room. Most mucus samples are currenly analysed by RT-PCR based methods for which the results do indicate a measure on the quantity of viral RNA. Since in an efficient PCR, a Ct value that is 9-10 cycli lower represents a virus RNA load that is 500-1000 higher, this information can be used to indicate a warning for those that may possibly shed much more than the average person. In the own data set the average detection is at Ct 26.1 cycli and about 2.5% shedders at Ct≤17 cycli were found, that can be indicated as probable suppershedders.

A superspreading event probably has multiple components. A person producing high levels of virus in the (upper) respiratory system can be thought of as a superreplicator. If a superreplicator is average or above average in droplet and aerosol production he or she may be a supershedder: not only producing high load of virus but also excreting them in transmittable form. If a supershedder does not follow the mitigation protocols (stay at home when sick, proper coughing and sneezing hygiene etc.) a supershedder can become a superspreader: someone causing many more infections than anticpated based on the average R_0_. The exposure assessment of this study demonstrates that viral RNA copy concentrations in mucus above 10^8^ per mL may easily give rise to very high exposure probabilities, even during breathing and speaking. SARS-CoV-2 superspreaders might be pre- or asymptomatic. Kimball et al. (2020) reported no significant difference in observed viral concentrations (Ct values) in throat/nose swabs between symptomatic, presymptomatic and asymptomatic individuals. One of the subjects in the study by Zou et al. (2020) was asymptomatic, and shed viral numbers that were similar to symptomatic subjects.

To which extent SARS-CoV-2 can be transmitted via aerosols has implications for intervention measures such as the advice to revert to community masking. Several meta-analyses have shown little protective effect to support widespread use of facemasks against COVID-19 (Brainard et al. 2020). However, there is enough evidence to support the use of facemasks for short periods of time by particularly vulnerable individuals when in transient higher risk situations.

The aim of this study was to assess exposure to airborne SARS-CoV-2 particles from breathing, speaking, coughing and sneezing in an indoor environment. Key findings are:

1. Data on size distribution, and amounts of aerosol droplets generated by breathing, speaking, coughing and sneezing that are small enough to stay afloat in the air and be dispersed in indoor spaces, vary widely. Despite this variation, there is ample evidence that indoor dispersion of aerosol droplets occurs. Generally, in more recent studies, smaller aerosol droplets, up to the submicron level, could be detected. Several studies have measured SARS-CoV-2 in the air, of which two showed infectious virus particles. Detection of viruses particles in the air is hampered by the fact that concentrations are commonly low, as was demonstrated in this study. Nevertheless, such concentrations may still give rise to significant probabilities of exposure (10 persons may inhale 5000 litres of air per hour). To conclude: aerosol transmission of SARS-CoV-2 is possible and should not be disregarded.
2. Moreover, according to the RIVM data, 20% of cases carry concentrations as high as at least 10^7^ RNA copies per mL mucus and 5% at least 10^8^ RNA copies per mL, probably even in pre- and asymptomatic persons, adding to the probability of exposure.
3. According to our results, sneezing leads to highest probabilities of exposure, followed by coughing and speaking and lastly breathing in the selected scenarios.
4. The exposure assessment in this study can be used as a basis to estimate probabilities of exposure to SARS-CoV-2 by airborne transmission in indoor spaces, thereby accounting for the probability that infected persons are present (prevalence), the number of exposed persons, the size of the indoor space, duration of expelling virus by the emitter, duration of exposure to the virus.
5. As long as it is uncertain what fraction of the airborne RNA copies relate to virus particles and how much of these are infectious and as long as a dose response relation is lacking, it is recommended to be precautious.
6. The exposure assessment could be improved by including heterogeneity of the (*eg*. directed) spreading of aerosol droplets, as well as its time dynamics.

## Data Availability

Data could be made available upon request.

## Acknowledgement

We gratefully acknowledge the Dutch COVID-19 response team existing of colleagues from RIVM-LCI, RIVM-EPI and Erasmus MC. Furthermore, we are grateful to the RIVM COVID-19 molecular diagnostic team of IDS for use of diagnostic data on Dutch COVID-19 patients. Municipal health services and hospital labs are thanked for sending in specimens of suspect COVID-19 patients and providing additional clinical data. Special thanks for Harry Vennema for fruitfull discussions. This work was funded by the Dutch Ministry of Health, Welfare, and Sports (VWS).

